# Longer Temporal Interference Stimulation Induces Detectable Intrinsic Activity Alterations in Human Nucleus Accumbens

**DOI:** 10.1101/2025.10.31.25339211

**Authors:** Zhenxiang Zang, Yongxi Zhang, Zhi Yang

## Abstract

The nucleus accumbens (NAc) possesses critical neurobiological and pathophysiological significance as a hub within the mesolimbic reward circuitry. Recent advances in non-invasive neuromodulation have demonstrated that temporal interference (TI) stimulation can achieve focal deep brain targeting in humans with favorable safety profiles. Therefore, TI stimulation on the NAc holds substantial promise for treating neuropsychiatric disorders. While stimulation duration represents an understudied parameter that may critically influence clinical outcomes, its impact on TI-induced NAc neuromodulation remains entirely unexplored. To address this knowledge gap, we employed a randomized, within-subject, counterbalanced crossover design in 24 healthy volunteers (HVs). Participants underwent both 20-minute or 40-minute 130 Hz TI stimulation targeting the NAc. Resting-state functional MRI (fMRI) data were acquired pre- and post-stimulation. Regional homogeneity (ReHo) and seed-based functional connectivity analyses were utilized to quantify intrinsic brain activity and connectivity. Paired t-tests revealed that 40-minute TI stimulation significantly attenuated NAc’ ReHo (P = 0.04), an effect not observed for 20 minutes of stimulation. Exploratory analyses revealed that the 40-minute stimulation also reduced ReHo of the orbitofrontal cortex (OFC) and attenuated NAc-OFC functional connectivity (Ps < 0.005). Critically, the 20-minute TI elicited no significant alterations. These findings demonstrate a duration-dependent response, where a longer (40-minute) TI protocol produced detectable functional alterations in the NAc and NAc-OFC pathway. Our study provides the first neuroimaging evidence that stimulation duration is a key parameter influencing TI-induced neuromodulatory effects, thereby providing crucial guidance for parameter selection in future clinical applications.

## Introduction

The nucleus accumbens (NAc), a subcortical structure located within the ventral striatum, serves as a critical integrative hub of the mesolimbic reward circuitry [1, 2]. Dysfunction of the NAc exhibits strong pathophysiological associations with diverse neuropsychiatric disorders, including depression, addiction, and obsessive-compulsive disorder [3-5]. Modulations of the aberrant NAc activity represent a promising therapeutic strategy for symptom relief [6-8].

Current approaches for precise NAc neuromodulation remain limited. Neurosurgical implantation of deep-brain stimulation (DBS) electrodes enables targeted modulation in treatment-resistant depression [9]. However, the invasive nature of DBS constrains its clinical applicability. While non-invasive alternatives such as transcranial magnetic stimulation (TMS) are widely employed, their limited stimulation depth precludes direct engagement of deep subcortical structures.

Temporal Interference (TI) is a novel electrical stimulation technique that enables precise modulation of deep brain regions. Briefly, TI utilizes intersecting high-frequency electric fields (e.g., 2 kHz and 2.1 kHz) to generate amplitude-modulated envelope frequencies (e.g., 100 Hz) at focal deep brain targets [10]. This approach enables spatially selective neuromodulation without activating superficial tissue. Previous studies have demonstrated TI’s capability to safely and effectively modulate hippocampal activity and enhance working memory [11]. In addition, TI’s frequency-specific stimulation over the striatum can influence motor learning processes in human [12, 13]. Clinical studies have further demonstrated that TI stimulation on the right internal globus pallidus (GPi) can significantly decrease motor dysfunction in patients with Parkinson’s disease [14]. Collectively, these findings establish TI as a capable strategy for precision engagement of deep brain structures. Therefore, NAc-targeted TI represents a promising therapeutic modality for neuropsychiatric disorders.

The translation of TI from neuroscientific discovery to clinical application necessitates rigorous parameter optimization. Beyond established parameters (e.g., envelope frequency or waveform morphology), stimulation duration remains an under-investigated variable with profound clinical implications. Existing evidence indicates that neuromodulatory outcomes exhibit temporal dependency, where differential exposure durations elicit distinct neurophysiological responses [15, 16]. This raises a fundamental question: Do TI-induced neuromodulatory effects on the NAc demonstrate duration-dependent neurobiological signatures?

To address this knowledge gap, we implemented a randomized, counterbalanced, within-subject crossover design in healthy volunteers (HVs). Participants received both 20-minute and 40-minute high-gamma TI stimulation targeting the NAc. Resting-state functional MRI (rs-fMRI) was acquired pre- and post-stimulation to quantify modifications of intrinsic brain activity and functional connectivity. Our primary objective was to characterize duration-dependent alterations in the NAc’s intrinsic activity and functional connectivity. This study seeks to provide the first neuroimaging evidence establishing stimulation duration as a critical determinant of TI efficacy, thereby informing parameter selection for future clinical trials.

## Materials and Methods

### Study Design and Participants

The current study was approved by the ethic committee of Beijing Anding Hospital. Twenty-four HVs (mean age = 27.45 years with 5.32 standard deviation, 12 females) participated in the current study after providing written informed consent. All participants were right-handed, free from any neurological or psychiatric disorders, and had no contraindications for MRI or electrical stimulation. We carried out a randomized, within-subject, counterbalanced cross-over design. Each subject received both 20-minute and 40-minute 130 Hz TI stimulation as did in DBS studies [17]. Twelve HVs were randomly assigned to 20-minute-first group and the rest were assigned to 40-minute-first group. In each session, participants immediately received TI stimulation after the baseline MRI scan. After receiving TI stimulation, all participants went back for follow-up scan immediately. A14-day interval was designed to wash out potential carry-over effect of TI based on prior pharmacological intervention study [18].

### MRI data acquisition and processing

All MRI data were acquired on a 3.0T GE Premier scanner with the following parameters: TR = 2000 ms, TE = 30 ms, number of slices = 50, interlaced axial scanning, slice thickness/gap = 3.5/0.7 mm; flip angle = 90°, matrix = 64 × 64; field of view (FOV) = 200 × 200 mm^2^, and voxel size = 3.13 × 3.13 × 4.2 mm^3^. T1 images were acquired using the magnetization-prepared rapid acquisition gradient echo sequence (MPRAGE): TR = 2530 ms, TE = 1.85 ms, number of sagittal slices = 192, slice thickness = 1 mm, FA = 15°; matrix = 256 × 256, FOV = 256 × 256 mm^2^, and voxel size = 1 × 1 × 1 mm^3^. During the resting-state fMRI data acquisition, all participants were instructed to fixate on a “+”. FMRI data were preprocessed using the SPM12, including the following steps: 1) head motion correction; 2) slice timing; 3) T1 registration and segmentation; 4) Indirect spatial normalization via T1 segmentations; 5) nuisance regression, including white mater, cerebral fluid signals, Power’s frame-wise displacement and head motion parameters and their squared parameters; 6) band-pass filtering of 0.01-0.1 Hz.

### Individualized TI stimulation

TI stimulation was achieved using the NervioX-1000 device provided by Xi’an NeuroDome Medical Technology Co., Ltd. An individualized TI stimulation strategy was applied to account for brain morphometry heterogeneity. The individualized TI stimulation was achieved as follow: 1) 3D-T1 images were segmented into six tissues upon SPM12 segmentation approach (http://www.fil.ion.ucl.ac.uk/spm/); 2) conductivities assignment to the scalp (2e-4 S/m), skull (0.0202 S/m), cerebrospinal fluid (2 S/m), white matter (0.064 S/m), gray matter (0.103 S/m), and cavity (2.5e-14 S/m) [19]; 3) registration of 10-10 EEG electrode system onto the participants’ scalp; 4) tetrahedral finite element meshes of the individual head model were generated using the open-source iso2mesh toolbox [20]; 5) computation of individual’s electric field intensity via finite element solver GetDP [21]; 6) establishing individualized TI electric field intensity using TI electric field coupling principle [10]; 7) identification of the anatomical location of NAc in MNI space based on Harvard-Oxford subcortical atlas and 8) the optimal electrode configuration was determined using convex optimization algorithms to maximize both electric field intensity and focal specificity [22]. Detailed TI parameters for each subject was obtained and shown in Table1. Before TI stimulation, the resistivity per unit is less than 8kΩ. The total current was 4 mA. Ramp-up and Ramp-down was 30 seconds.

**Table 1.**
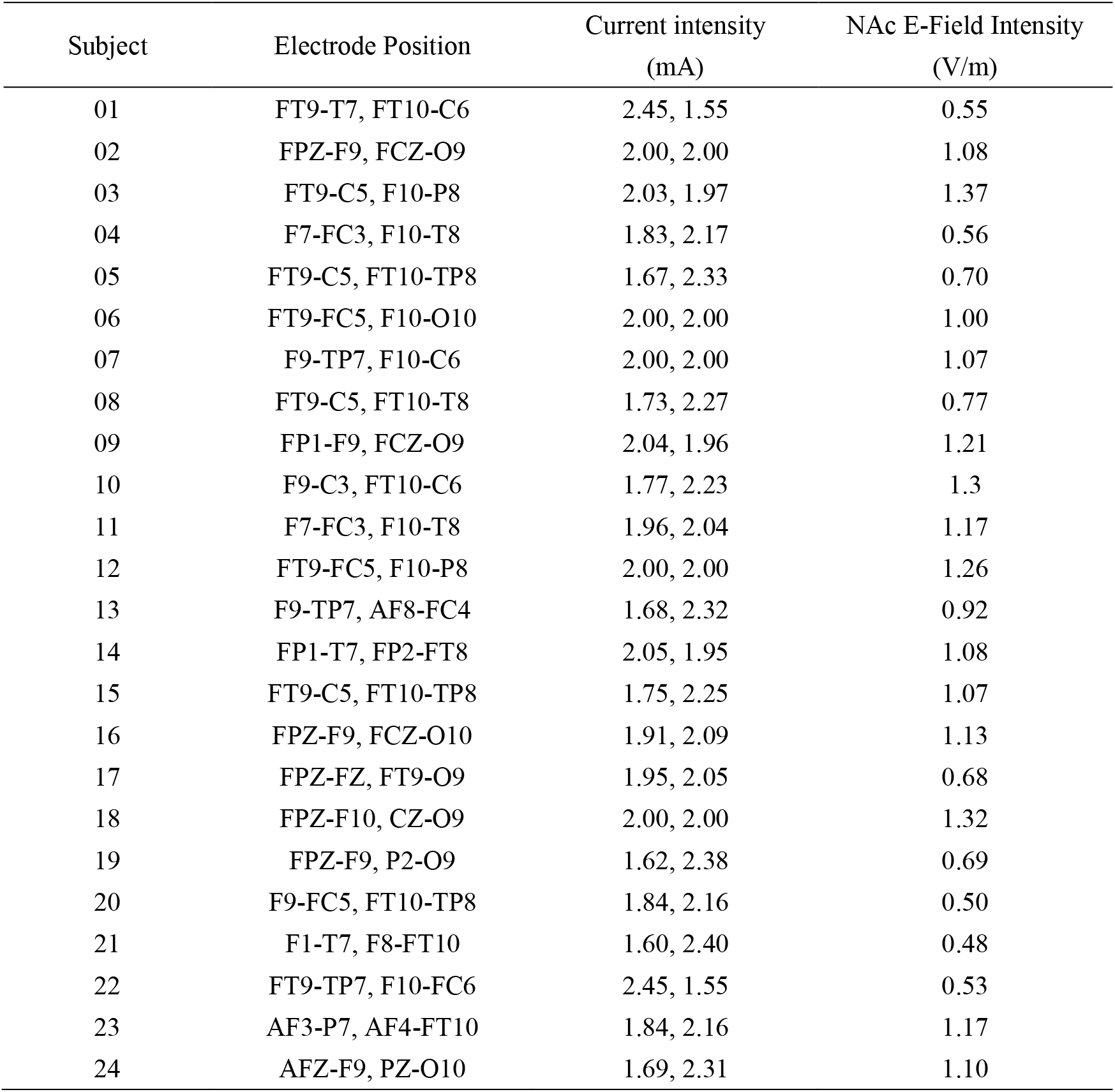
Electrode position, current intensity (mA), and estimated electric field intensity of the NAc (V/m, Harvard-Oxford subcortical) for each subject.

### Brain functional metrics

We used Regional Homogeneity (ReHo), a measurement of local synchrony, calculated as the temporal similarity (Kendall’s coefficient) of a voxel with its neighbors’ 27-voxel [23]. ReHo was spatially normalized by dividing by the global mean. We also calculate functional connectivity (FC) with Pearson’s correlation coefficient between clusters or regions showing a significant effect of TI stimulation to measure the inter-region connectivity. Both ReHo and FC were then smoothed (6-mm kernel).

### Statistical Analyses

Our main analysis followed a two-layer (hypothesis-driven and exploratory) framework to identify the effect of stimulation duration on NAc. First, we extracted the averaged ReHo values from the Harvard-Oxford subcortical NAc ROI and performed a 2 (condition: 20-minute, 40-minute) × 2 (time: pre, post) repeated-measures ANOVA (rmANOVA) as well as paired t-tests to compare the different effects of stimulation duration on the NAc. To investigate the spatial specificity of the NAc, we also compared the effect of TI on the other subcortical regions, including the putamen, caudate, amygdala, hippocampus, globus pallidus and the thalamus using paired t-tests. We also counted the percentage of volume surviving P < 0.05 uncorrected threshold at the voxel-level within these subcortical regions. For these analyses, we hypothesized that only NAc responds to TI stimulation.

We further carried out exploratory analyses to obtain the TI-induced alterations in the cortex. For exploratory whole-brain analyses, we report clusters that survived GRF correction (e.g., cluster-level p-FWE < 0.05). To identify potential areas of interest for future studies, we also describe the location of effects observed at an uncorrected threshold (e.g., voxel p < 0.005, k > 20) [24], but explicitly note that these did not survive whole-brain correction.

## 3. Results

### 3.1 Electric Field Modeling Validates the NAc as the Primary Stimulation Target

For visualization purpose, we first transformed the individual TI electric field map into the MNI space and calculated the mean simulated electric field intensity. The mean electric field intensity for the NAc is 0.95 ± 0.29 V/m and ranks on the top of all regions. For cortical areas (extracted from Schaefer’s template [25]), the subgeneual anterior cingulate cortex and the orbital frontal cortex (OFC) ranked on the top, with mean electric field intensity greater than 0.8 V/m. The distribution of the simulated electric field intensity is shown in Figure 1.

**Figure 1.**
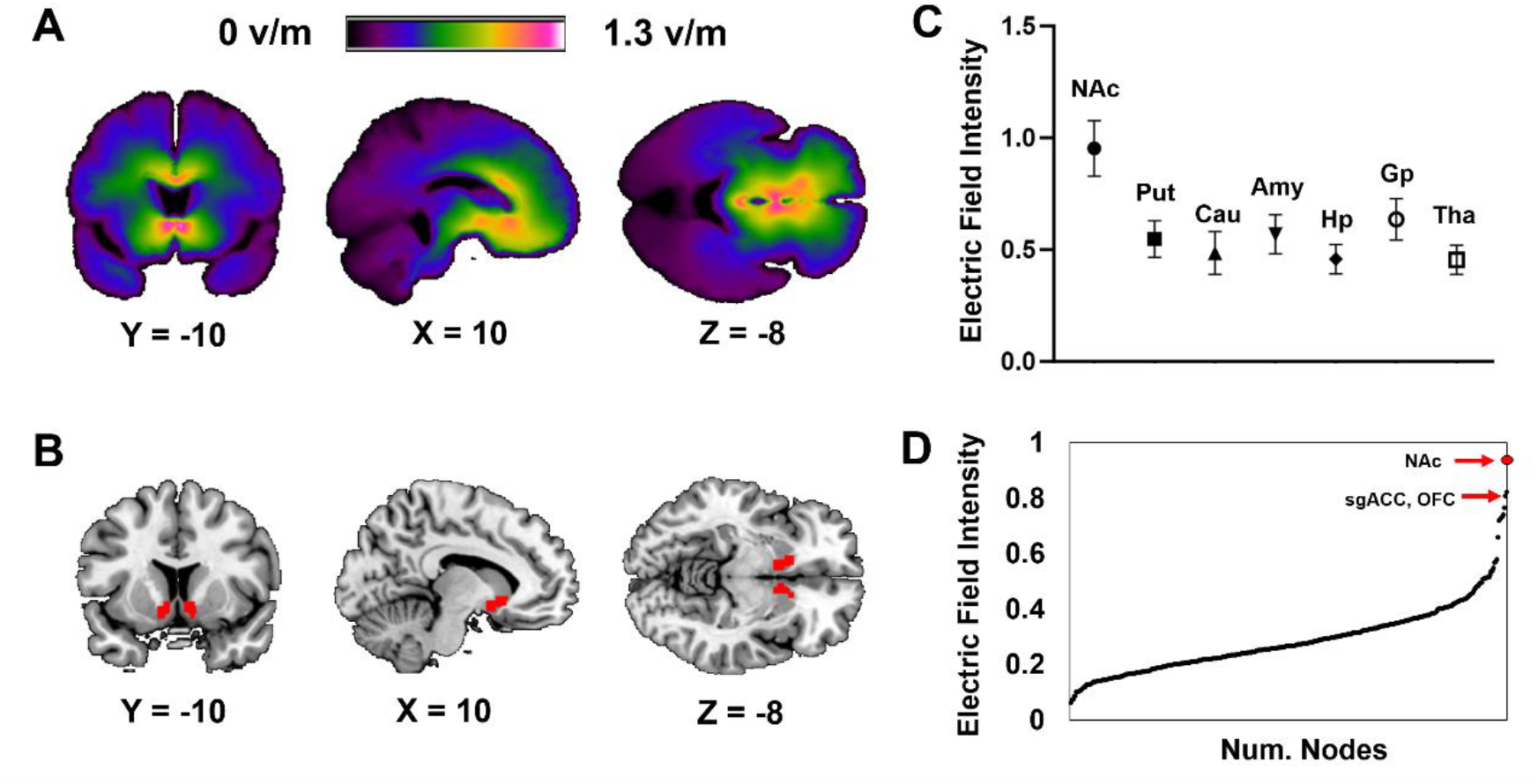
Electric strength distribution of the brain. (A) Whole-brain mean electric intensity distribution of all subjects in the MNI space. (B) Location of NAc template. (C) Distribution of the electric field intensity among subcortical regions, error bars indicate standard deviations. (D) Distribution of the electric field intensity among subcortical regions extracted from Schaefer’s 400 template. Abbreviations: NAc, nucleus accumbens; Put, putamen; Cau, caudate; Amy, amygdala; Hp, hippocampus; Gp, globus pallidus; Tha, thalamus; sgACC, subgenual anterior cingulate cortex; OFC, orbital frontal cortex.

### 3.2 Longer Stimulation Duration Preferentially Attenuates NAc Intrinsic Activity

Our primary hypothesis was that the neuromodulatory effect of TI on the NAc would be dependent on stimulation duration. A 2 (condition: 20-minute, 40-minute) × 2 (time: pre-, post-stimulation) repeated-measures ANOVA on NAc’s ReHo revealed a strong trend towards a significant condition-by-time interaction (F_1, 23_ = 4.15, P = 0.053, η^2^ = 0.15), indicating that the change in ReHo differed between the two protocols (Figure 2A).

**Figure 2.**
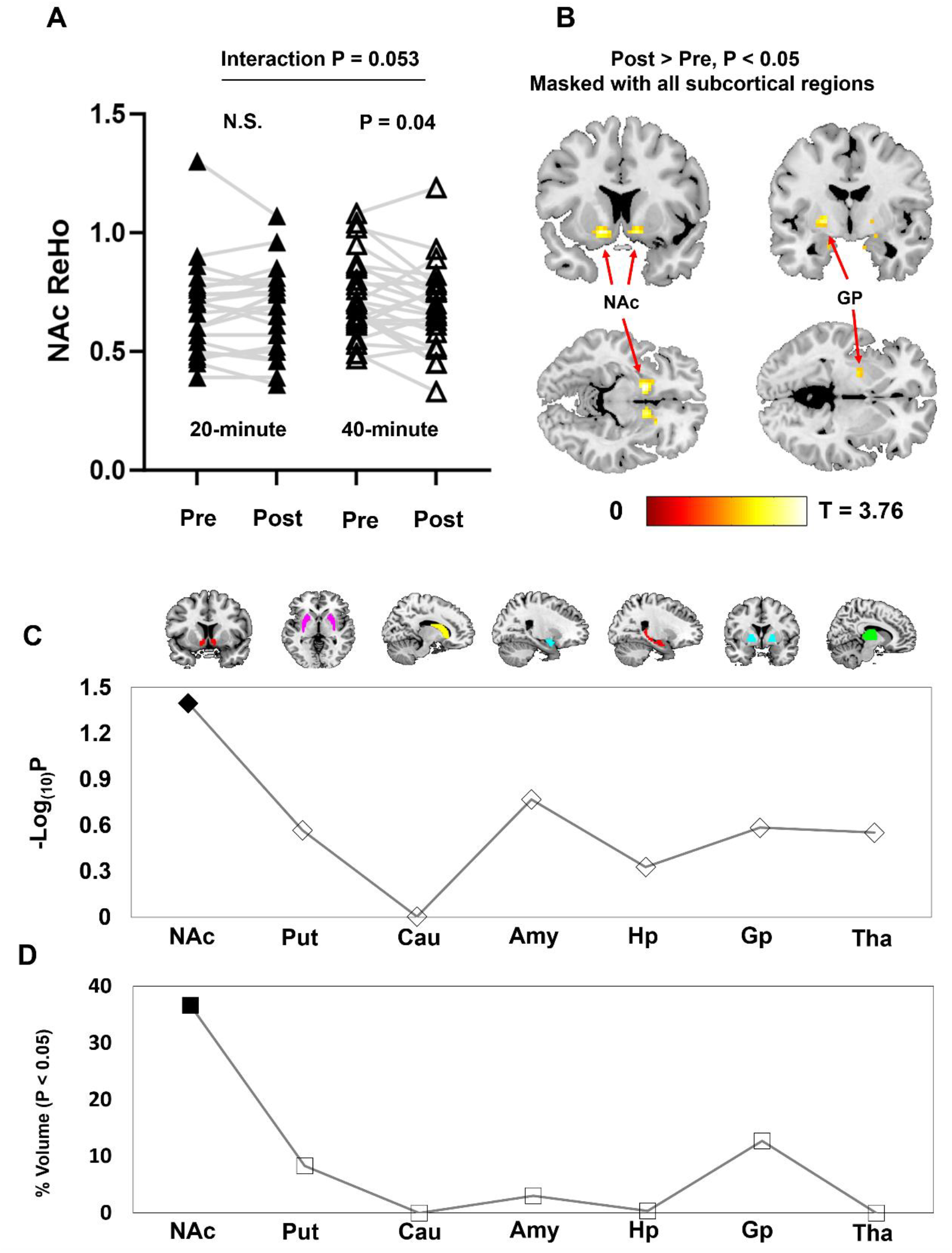
NAc and other subcortical ReHo response to 20-minute and 40-minute TI stimulation. (A) 40-minute TI stimulation significantly decreased NAc ReHo (P = 0.04) while 20-minute TI did not, resulting in marginal interaction effect (P = 0.053). (B) Spatial distribution of the voxels with P < 0.05 uncorrected threshold in all subcortical regions from the Harvard-Oxford template. Except for NAc, a cluster with 20 voxels adjacent to the globus pallidus (GP) can also be visualized showing reduced ReHo after 40-minute TI. No voxels showed increased ReHo within the NAc. (C) NAc is the only region showing significant reduced ReHo after 40-minute TI stimulation among the sub-cortical regions. (D) NAc’s percentage of volume (num. voxels / total voxels) that survived P < 0.05 uncorrected threshold ranked on the top among sub-cortical regions, and is larger than the sum of the rest (36.7% v.s. 24.5%). Abbreviations: NAc, nucleus accumbens; Put, putamen; Cau, caudate; Amy, amygdala; Hp, hippocampus; Gp, globus pallidus; Tha, thalamus.

To decompose this interaction, follow-up paired t-tests were performed. The results confirmed that the 40-minute TI protocol induced a significant reduction in NAc’s ReHo (T_23_ = 2.15, P = 0.04, 95% CI = [0.002 – 0.109]). In contrast, the 20-minute TI protocol elicited no significant change in NAc’s ReHo (T_23_ = 0.23, P = 0.79, 95% CI = [-0.041 – 0.032]). We did not obtain any significant alterations in other subcortical regions (all P > 0.17). In addition, we explored the subcortical ReHo alterations by using a P < 0.05 uncorrected threshold and obtained that the attenuation effect was perfectly distributed in the NAc at voxel-level (Figure 2B). There were nearly 37% of the volume survived P < 0.05 uncorrected threshold in the NAc, which was larger than the other subcortical regions (Figure 2C-D).

Control analyses confirmed the robustness of these findings. There was no significant effect of stimulation order (P > 0.18), nor significant baseline difference of NAc (P > 0.08). Furthermore, the magnitude of the ReHo change in the NAc did not correlate with the subject-specific simulated electric field intensity for either stimulation duration (all P > 0.55).

### 3.3 Exploratory Analysis Reveals Duration-Dependent Modulation of the OFC and NAc-OFC Connectivity

To investigate TI-induced effects beyond our primary subcortical target, we conducted an exploratory whole-brain analysis of ReHo changes. This analysis revealed no cortical regions with significant effects that survived GRF correction for multiple comparisons.

However, to identify potential regions of interest for future studies, we examined effects at an uncorrected threshold (voxel-wise P < 0.005, cluster size > 20 voxels). This exploratory step identified two clusters in the bilateral orbitofrontal cortex (OFC; Brodmann Area 47) that showed reduced ReHo exclusively after 40 minutes of TI stimulation (Figure 3A). The first cluster was located in the left OFC (40 voxels; peak MNI: -24, 21, -15; T_23_ = 3.93), and the second in the right OFC (28 voxels; peak MNI: 30, 24, -15; T_23_ = 4.88). Direct comparison confirmed this duration-dependency, with both the left and right OFC clusters demonstrating a significant condition-by-time interaction (left: F_1, 23_ = 15.94, P < 0.001, η^2^ = 0.41; right: F_1, 23_ = 11.91, P = 0.002, η^2^ = 0.34), driven by the lack of change in the 20-minute condition (Figure 3B-C).

**Figure 3.**
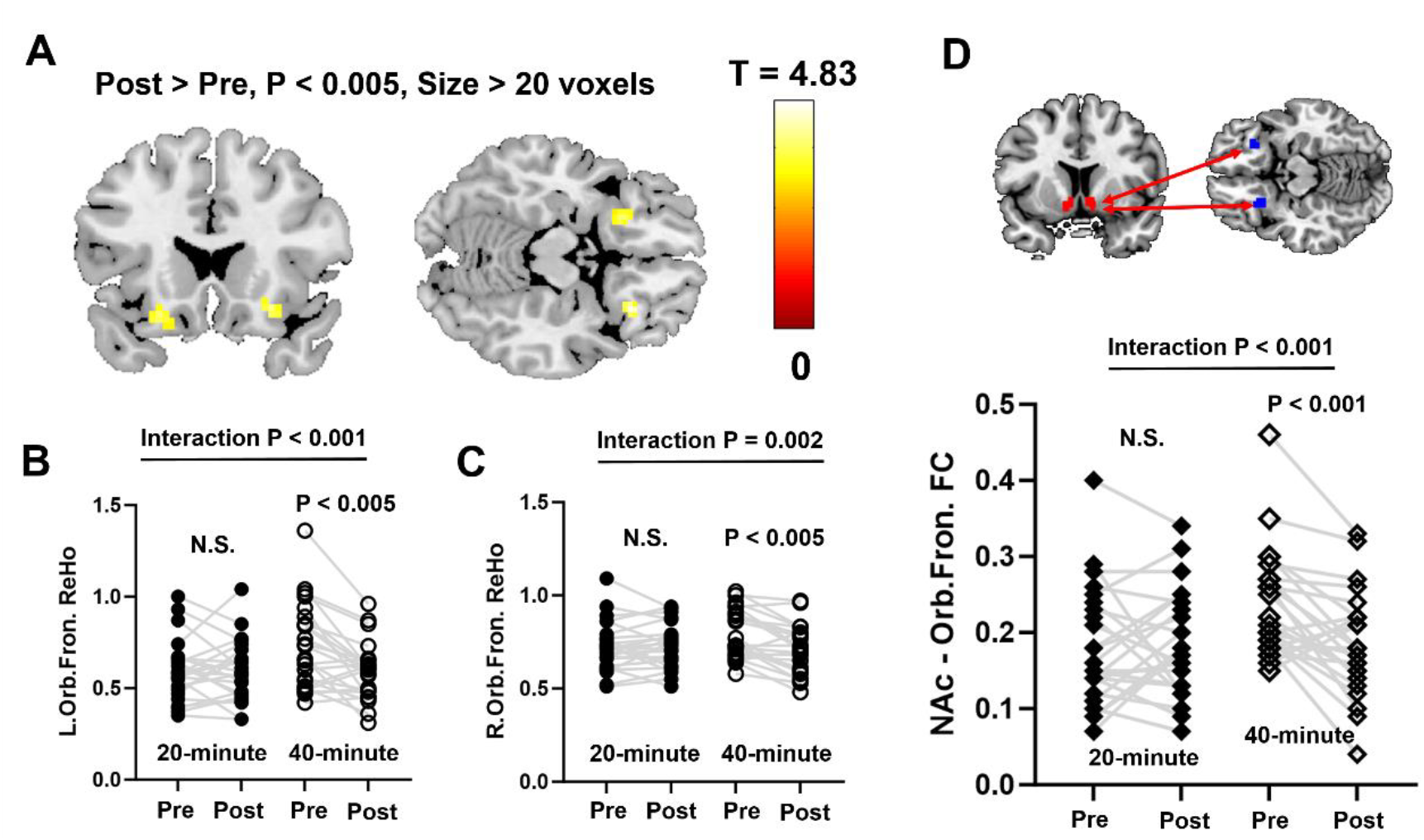
Exploratory results of 40-minute TI stimulation’s effect in the orbital frontal cortex. (A) Two clusters (uncorrected P < 0.005, size > 20 voxels) were found in the bilateral orbital frontal cortex with a marginal significant 40-minute TI attenuation effect. (B) and (C), These orbital frontal clusters did not show significant alterations for 20-minute TI stimulation, resulting in significant condition-by-time interactions. (D) 40-minute TI, but not 20-minute TI, also induced a reduction FC between the NAc and the bilateral OFC clusters.

Given the established connectivity between the NAc and OFC, we then tested whether TI modulated this pathway. Using the bilateral OFC clusters as a functional region of interest, we found that NAc-OFC functional connectivity was also significantly reduced following the 40-minute TI protocol (T_23_ = 3.97, P < 0.001), but not the 20-minute protocol. This resulted in a robust condition-by-time interaction (F_1, 23_ = 14.23, P < 0.001, η^2^ = 0.38; Figure 3D). Finally, neither the ReHo changes in the OFC nor the NAc-OFC connectivity changes were correlated with the simulated electric field intensity within the OFC (all P > 0.29).

## Discussion

The NAc stands as a pivotal therapeutic target for a range of severe neuropsychiatric disorders, yet modulating this deep subcortical structure non-invasively has remained a major clinical challenge. Addressing this critical gap, the current study investigated whether TI stimulation, a novel non-invasive technique, could be optimized to engage the NAc by systematically varying stimulation duration. Utilizing a rigorous, randomized within-subject design and individualized electric field modeling, we provide the first empirical evidence for a duration-dependent neuromodulatory effect of TI on the human brain. Our results clearly demonstrate that a longer (40-minute) stimulation protocol successfully attenuated intrinsic neural activity in the NAc and its functionally connected orbitofrontal pathway—an effect that was absent following a shorter 20-minute protocol. This work validates that individualized TI can precisely engage a small, deep subcortical nucleus, but more critically, it establishes stimulation duration as a key parameter influencing the efficacy of neuromodulation. These findings offer crucial guidance for optimizing TI protocols, moving this promising therapeutic tool a substantive step closer to clinical application.

Previous TI research has predominantly focused on stimulation efficacy (e.g., on/off effects) and frequency-specific modulation. For instance, Violante et al. reported that 5-Hz reduced hippocampal activation during working memory tasks [11]. Vassiliadis et al., found that 80-Hz, not 20-Hz TI stimulation on the striatum could disrupt motor learning [12]. While these studies demonstrated TI’s capacity to modulate deep brain regions, the critical parameter of stimulation duration remains largely unexplored. In addition, the effect of stimulation duration cannot be directly tested using on/off experimental design. Although duration may not be the primary determinant of TI’s neurobiological effects, it likely represents a crucial parameter influencing clinical translation. As an example, Nettekoven et al. demonstrated a dose-dependent effect of theta-burst transcranial magnetic stimulation, where extended protocols (1800 pulses), but not brief ones, significantly enhanced functional connectivity [26]. Consistent with this finding, our data revealed that 40-minute, but not 20-minute TI stimulation, produced a significant attenuation effect of NAc intrinsic activity, suggesting a potential preference for longer protocols.

Prior TI neuroimaging studies primarily targeted larger subcortical structures. While some reported successful target engagement, others failed to detect significant changes in regions like the caudate [27]. These studies typically leveraged simulated electric fields based on standard templates; however, inherent complexities in brain tissue conductivity can lead to substantial spatial heterogeneity in actual field distributions. Liu et al. recently quantified the spatial correlation between simulated and empirically measured electric fields in primates, revealing non-negligible inter-subject variability [28]. Such anatomical and biophysical heterogeneity likely amplifies targeting uncertainty, particularly for small, deep nuclei like the NAc. To avoid this, we employed individualized electric field modeling. As expected, although strikingly, results highlighted the NAc as the subcortical region with the most robust and statistically significant functional modulation, underscoring the utility of subject-specific field modeling for precision targeting of TI.

Furthermore, we observed significantly reduced functional connectivity between the NAc and OFC, alongside a marginal decreased OFC intrinsic activity, following 40-minute (versus 20-minute) TI. This demonstrates that the duration-dependent effect may extend to the NAc-OFC pathway. While this finding is anatomically plausible, given the established structural connectivity between NAc and OFC [29] and their functional integration in affective regulation and reward processing [30, 31], it must be interpreted with caution. The OFC itself received a substantial electric field, and the statistical evidence for this effect did not survive correction for whole-brain multiple comparisons. Therefore, it remains unclear whether this reflects a downstream effect of NAc modulation or a parallel, direct stimulation of the OFC. We did not observe a direct correlation between the simulated E-field intensity in the NAc or OFC and the magnitude of the corresponding fMRI changes. This lack of a linear relationship may indicate a non-monotonic or threshold-based biological response, where a sufficient E-field is necessary to initiate a change, but supra-threshold variations do not linearly scale the effect.

This study has several limitations. First, our investigation focused on the effects of stimulation duration within a single session. Clinical applications, however, typically involve multiple-session treatment protocols. Cumulative session number may exert greater influence on treatment outcomes than per-session duration. Second, it is crucial not to assume a simple linear dose-response relationship where “more is better”. Robust evidence indicates that neurostimulation dose-treatment response relationships are frequently non-monotonic [32, 33]. Given these considerations, we deliberately constrained our investigation to durations ≤ 40 minutes. TI’s dose-dependent therapeutic efficacy requires demonstration from future clinical trials.

In conclusion, this study provides the first neuroimaging evidence that the neuromodulatory impact of TI is critically dependent on its stimulation duration. We demonstrated that a 40-minute protocol successfully attenuated intrinsic activity in the NAc and its functional connected OFC circuitry. This work not only substantiates the precision and efficacy of individualized TI for targeted deep brain neuromodulation but also provides empirical guidance for optimizing stimulation parameters (e.g., duration) towards developing NAc-targeted TI interventions for neuropsychiatric disorders.

## Data Availability

The raw MRI data, according to the local ethics committees regulation, is not allowed for distribution.

## CRediT authorship contribution statement

Zhenxiang Zang: Writing – review & editing, Writing – original draft, Visualization, Validation, Methodology, Investigation, Formal analysis, Conceptualization Yongxi Zhang: Writing – review & editing, Investigation, Data curation Zhi Yang: Writing – review & editing, Writing – original draft, Visualization, Validation, Methodology, Investigation, Conceptualization

## Acknowledgement

We thank Liwen Feng and Dr. Tian Liu for their effort in personalized localization of TI. We would also like to thank Xi’an NeuroDome Medical Technology Co., Ltd for providing TI device.

## AI-supported writing

During the preparation of this work the authors used ChatGPT in order to improve the readability. After using this service, the authors reviewed and edited the content as needed and take full responsibility for the content of the publication

## Data availability

The raw MRI data, according to the local ethics committee’s regulation, is not allowed for distribution without restriction. Processed data is accessible after obtaining agreement from the corresponding authors.

## References

[1] Salgado S, Kaplitt MG. The Nucleus Accumbens: A Comprehensive Review. Stereotact Funct Neurosurg 2015;93(2):75–93.

[2] Floresco SB. The nucleus accumbens: an interface between cognition, emotion, and action. Annu Rev Psychol 2015;66:25–52.

[3] Shirayama Y, Chaki S. Neurochemistry of the nucleus accumbens and its relevance to depression and antidepressant action in rodents. Curr Neuropharmacol 2006;4(4):277–91.

[4] Scofield MD, Heinsbroek JA, Gipson CD, Kupchik YM, Spencer S, Smith AC, et al. The Nucleus Accumbens: Mechanisms of Addiction across Drug Classes Reflect the Importance of Glutamate Homeostasis. Pharmacol Rev 2016;68(3):816–71.

[5] Xu L, Nan J, Lan Y. The Nucleus Accumbens: A Common Target in the Comorbidity of Depression and Addiction. Front Neural Circuits 2020;14:37.

[6] Bewernick BH, Hurlemann R, Matusch A, Kayser S, Grubert C, Hadrysiewicz B, et al. Nucleus accumbens deep brain stimulation decreases ratings of depression and anxiety in treatment-resistant depression. Biol Psychiatry 2010;67(2):110–6.

[7] Bewernick BH, Kayser S, Sturm V, Schlaepfer TE. Long-term effects of nucleus accumbens deep brain stimulation in treatment-resistant depression: evidence for sustained efficacy. Neuropsychopharmacology 2012;37(9):1975–85.

[8] Denys D, Mantione M, Figee M, van den Munckhof P, Koerselman F, Westenberg H, et al. Deep Brain Stimulation of the Nucleus Accumbens for Treatment-Refractory Obsessive-Compulsive Disorder. Archives of General Psychiatry 2010;67(10):1061–8.

[9] Shivacharan RS, Rolle CE, Barbosa DAN, Cunningham TN, Feng A, Johnson ND, et al. Pilot study of responsive nucleus accumbens deep brain stimulation for loss-of-control eating. Nature Medicine 2022;28(9):1791–6.

[10] Grossman N, Bono D, Dedic N, Kodandaramaiah SB, Rudenko A, Suk HJ, et al. Noninvasive Deep Brain Stimulation via Temporally Interfering Electric Fields. Cell 2017;169(6):1029-41.e16.

[11] Violante IR, Alania K, Cassarà AM, Neufeld E, Acerbo E, Carron R, et al. Non-invasive temporal interference electrical stimulation of the human hippocampus. Nat Neurosci 2023;26(11):1994–2004.

[12] Vassiliadis P, Beanato E, Popa T, Windel F, Morishita T, Neufeld E, et al. Non-invasive stimulation of the human striatum disrupts reinforcement learning of motor skills. Nat Hum Behav 2024;8(8):1581–98.

[13] Wessel MJ, Beanato E, Popa T, Windel F, Vassiliadis P, Menoud P, et al. Noninvasive theta-burst stimulation of the human striatum enhances striatal activity and motor skill learning. Nat Neurosci 2023;26(11):2005–16.

[14] Yang C, Xu Y, Feng X, Wang B, Du Y, Wang K, et al. Transcranial Temporal Interference Stimulation of the Right Globus Pallidus in Parkinson’s Disease. Mov Disord 2025;40(6):1061–9.

[15] Vasquez A, Malavera A, Doruk D, Morales-Quezada L, Carvalho S, Leite J, et al. Duration Dependent Effects of Transcranial Pulsed Current Stimulation (tPCS) Indexed by Electroencephalography. Neuromodulation: Technology at the Neural Interface 2016;19(7):679–88.

[16] Hutton TM, Aaronson ST, Carpenter LL, Pages K, Krantz D, Lucas L, et al. Dosing transcranial magnetic stimulation in major depressive disorder: Relations between number of treatment sessions and effectiveness in a large patient registry. Brain Stimulation: Basic, Translational, and Clinical Research in Neuromodulation 2023;16(5):1510–21.

[17] Dandekar MP, Fenoy AJ, Carvalho AF, Soares JC, Quevedo J. Deep brain stimulation for treatment-resistant depression: an integrative review of preclinical and clinical findings and translational implications. Molecular Psychiatry 2018;23(5):1094–112.

[18] Diazgranados N, Ibrahim L, Brutsche NE, Newberg A, Kronstein P, Khalife S, et al. A randomized add-on trial of an N-methyl-D-aspartate antagonist in treatment-resistant bipolar depression. Arch Gen Psychiatry 2010;67(8):793–802.

[19] Gabriel C. Compilation of the dielectric properties of body tissues at RF and microwave frequencies. Final Report for the Period 15 December 1994 -14 December 1995. In: Air Force Office of Scientific R, ed. US; 1996:1–272.

[20] Qianqian F, Boas DA. Tetrahedral mesh generation from volumetric binary and grayscale images. 2009 IEEE International Symposium on Biomedical Imaging: From Nano to Macro. 2009:1142–5.

[21] Geuzaine C. GetDP: a general finite-element solver for the de Rham complex[C]//PAMM: Proceedings in Applied Mathematics and Mechanics. Berlin: WILEY - VCH Verlag 2007;7(1):1010603–4.

[22] Geng C, Li Y, Li L, Zhu X, Hou X, Liu T. Optimized Temporal Interference Stimulation Based on Convex Optimization: A Computational Study. IEEE Trans Neural Syst Rehabil Eng 2025;33:1400–10.

[23] Zang Y, Jiang T, Lu Y, He Y, Tian L. Regional homogeneity approach to fMRI data analysis. Neuroimage 2004;22(1):394–400.

[24] Deininger-Czermak E, Spencer L, Zoelch N, Sankar A, Gascho D, Guggenberger R, et al. Magnetic resonance imaging of regional gray matter volume in persons who died by suicide. Molecular Psychiatry 2025;30(3):1029–33.

[25] Schaefer A, Kong R, Gordon EM, Laumann TO, Zuo XN, Holmes AJ, et al. Local-Global Parcellation of the Human Cerebral Cortex from Intrinsic Functional Connectivity MRI. Cereb Cortex 2018;28(9):3095–114.

[26] Nettekoven C, Volz LJ, Kutscha M, Pool EM, Rehme AK, Eickhoff SB, et al. Dose-dependent effects of theta burst rTMS on cortical excitability and resting-state connectivity of the human motor system. J Neurosci 2014;34(20):6849–59.

[27] Modak P, Fine J, Colon B, Need E, Cheng H, Hulvershorn L, et al. Temporal interference electrical neurostimulation at 20 Hz beat frequency leads to increased fMRI BOLD activation in orbitofrontal cortex in humans. Brain Stimul 2024;17(4):867–75.

[28] Liu R, Zhu G, Wu Z, Gan Y, Zhang J, Liu J, et al. Temporal interference stimulation targets deep primate brain. Neuroimage 2024;291:120581.

[29] Rusche T, Kaufmann J, Voges J. Nucleus accumbens projections: Validity and reliability of fiber reconstructions based on high-resolution diffusion-weighted MRI. Hum Brain Mapp 2021;42(18):5888–910.

[30] Winstanley CA, Theobald DE, Cardinal RN, Robbins TW. Contrasting roles of basolateral amygdala and orbitofrontal cortex in impulsive choice. J Neurosci 2004;24(20):4718–22.

[31] Catani M, Dell’Acqua F, Thiebaut de Schotten M. A revised limbic system model for memory, emotion and behaviour. Neuroscience & Biobehavioral Reviews 2013;37(8):1724–37.

[32] Batsikadze G, Moliadze V, Paulus W, Kuo MF, Nitsche MA. Partially non-linear stimulation intensity-dependent effects of direct current stimulation on motor cortex excitability in humans. J Physiol 2013;591(7):1987–2000.

[33] Sabé M, Hyde J, Cramer C, Eberhard A, Crippa A, Brunoni AR, et al. Transcranial Magnetic Stimulation and Transcranial Direct Current Stimulation Across Mental Disorders: A Systematic Review and Dose-Response Meta-Analysis. JAMA Netw Open 2024;7(5):e2412616.

